# Neuropathological Features of Fatal Electrocution: A Histopathological Study for Cause-of-Death determination

**DOI:** 10.64898/2026.09.21.26363180

**Authors:** Ashok k Rastogi, Ashutosh Kumar, Dinil D Nangelil, Sanjeev K Paikra, Mrunali D Dhokne, Yashwant M Reddy, Toshal d Wankhade, Rajesh Kumar

## Abstract

**Background:** Determining the cause of death in electrocution is often challenging at autopsy, particularly when characteristic external electrical marks are absent. Reliable internal markers are scarce, and the neuropathological changes associated with electrocution remain underexplored.

**Objective:** To identify and characterize histopathological alterations in the brain in cases of fatal electrocution and evaluate their diagnostic utility.

**Methods:** A prospective observational study was conducted on 12 autopsy cases, including 7 confirmed electrocution deaths and 5 non-electrocution deaths (control). Brain tissue samples from multiple regions were processed using H&E and Golgi–Cox staining. Histopathological parameters, including neuronal morphology, vascular changes, haemorrhage, and dendritic architecture, were evaluated.

**Results:** Electrocution cases demonstrated neuronal destruction, axonal ballooning, focal hemorrhages, and dendritic disruption. Cerebellar involvement with Purkinje cell damage.

**Conclusion:** Comprehensive neurohistological technique may helpful in evaluation neuronal damage in electrocution death. Neuropathological findings, particularly neuronal and dendritic injury, may serve as supportive evidence in determining cause of death when external signs are absent.

## Introduction

The determination of death due to electrocution depends on the morphological observations of the electric mark (1). About 20-25% electrocution deaths occur without detectable current marks on the skin (2), making a forensic determination of the cause of death more difficult (3–5), especially when it passes through the moist surface of the body.At our autopsy centre, electrocution deaths accounted for 3% of the total autopsies conducted over the last two years. In forensic practice, establishing electrocution as the cause of death can be challenging, especially when classical external findings such as entry and exit burns mark absent.

Lacking of comprehensive neuronal injury evaluation in determination of cause of death in electrocution. Histopathological examination offers a potential adjunct in such cases, this study aims to evaluate neuropathological changes in fatal electrocution and assess their diagnostic relevance. The integration of these techniques may enhance understanding of electrical injury to the central nervous system.

## Materials and Methods

Brain tissue samples harvested during autopsy, 3 sample collectected in each autopsy case, from temporal, parietal and cerebellum part of brain, one sample from each lobe. Total 12 autopsy cases were taken, 7 test cases (electrocution deaths) and 5 control cases (non-electrocution deaths).

The cause of death was confirmed by the autopsy surgeon in all cases. Convenient sampling was performed after obtaining written informed consent from the kin. Sample of size approx 2 × 2 cm were excised and preserved.

### H& E Staining

The sample were fixed in 4% paraformaldehyde (PFA) for 6 hours, then transferred to 30% sucrose, kept in 4 degree sentigrade temperature until the sectioning process of the sample.

### Golgi staining

Tissue impregnation done in Golgi-Cox solution was prepared by dissolving 16 g (6% w/v) each of the following chemicals in 300 mL distilled water (dH2O): Potassium dichromate (K2Cr2O7), Mercuric chloride (HgCl2) and Potassium chromate (K2CrO4).

The all three stock solutions were stored in separate bottles at room temperature in the dark for use in preparing Golgi-Cox solution. For each experiment, fresh Golgi-Cox solution was prepared in a new bottle using the stock solutions as follows (6). 60 mL potassium dichromate solution was mixed with 60 mL mercuric chloride solution. 40 mL potassium chromate solution was added and 100 mL double-distilled water (dH2O) was added. Samples were kept in Golgi cox solution for 10 days.

(After mixing the solution, the bottle needs to be covered with aluminum foil and kept at room temperature for at least 48 h before use to allow precipitate formation. This solution may be for up to 2 month.)

### Sectioning of Samples

Brain samples were embedded in 4% low-melting-point agarose by pouring the solution into a mold. The sample position was adjusted using a pipet tip so that the cut surface faced downward. The agarose was allowed to fully harden.

Once solidified, the mold edges and sides were trimmed, and excess agarose was removed with a razor blade. The trimmed agarose block was mounted onto the vibratome plate and placed in the chamber, which was filled with tissue-protectant solution until the block was fully covered.

Sections of 200 μm thickness were collected using a thick brush and transferred onto 4 % gelatin-coated slides.

### Developing steps

The tissue sections were serially passed through the developing solutions (approximately 300 mL of each) (Box 1) for fixed times at room temperature.

#### Box-1

Steps and solutions for the development of neuronal structure/ arborisation.

- Xylene (C8H10): 6 min × 2
- Ethanol series C2H6O: 60%, 70%, 96%, 100% (6 min each)
- 3:1 ammonia:dH2O (prepared by mixing 200 mL ammonia (NH3H3O): 10 min
- 6% sodium thiosulfate (prepared by dissolving 16 g sodium thiosulfate (Na2S2O3H2O) in 300 mL dH2O: 8 min
- Distilled water: 1min
- Xylene: 6 min x 2 (for dehydration and clearing)

After staining slides air-dried then mounted, all solutions were stored at room temperature, the sodium thiosulfate bottle was covered with aluminum foil. Solutions could be reused and were replaced when they turned hezy/dark. Structural changes were assessed in the first sample using hematoxylin and eosin (H&E) staining and stained section were observed under a light microscope.

**Table 1:** Details of entry and exit wound in electrocution deaths cases.

| S.N. | Age group (years) /Sex | TSD | Entry wound | Exit wound |
| --- | --- | --- | --- | --- |
| 1. | 25-30/M | 20hr | Right hand | left hand |
| 2. | 35-40/M | 18hr | Left hand | Left foot |
| 3. | 30-35/M | 15hr | Left side head | Right hand |
| 4. | 40-45/M | 18hrs | Left hand | Left foot |
| 5. | 50-55/M | 15hrs | Right hand | Left hand |
| 6. | 20-25/M | 9hrs | Right side head | Right foot toe |
| 7. | 5-10/M | 6hrs | Right foot | Right hand |

## Results

### Macroscopic observations

Samples were grossly examined during autopsy, we found multiple petechial haemorrhages in white matter, as Shown in Fig. 01. In the grey matter by the naked eye after fixation of the tissue, no any remarkable observations.

**Figure 01:**
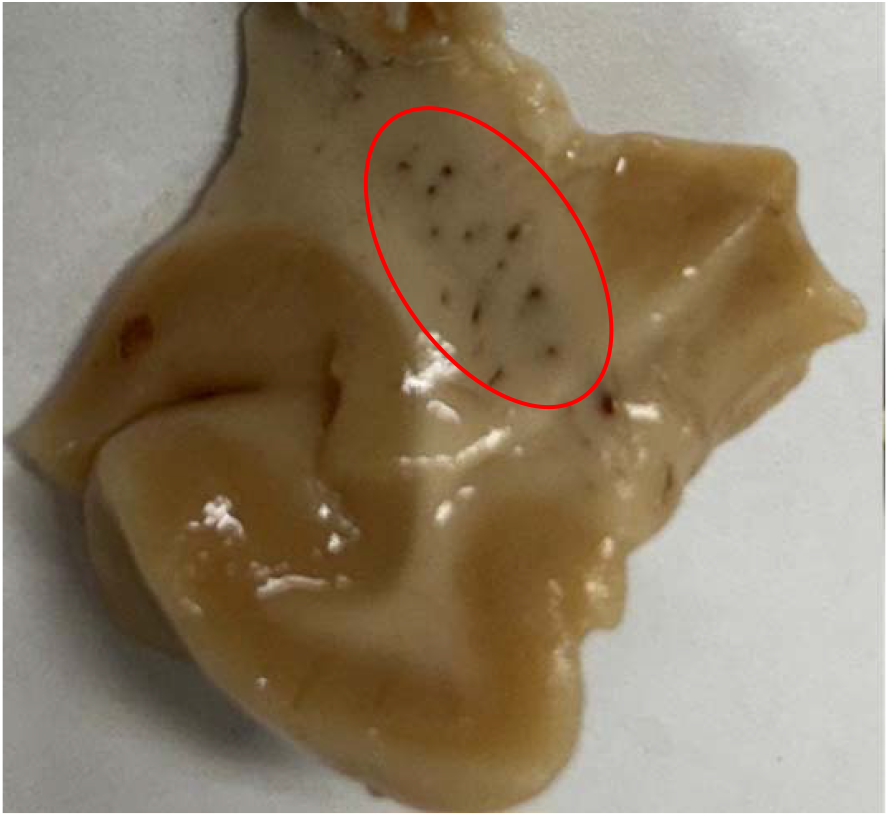
Observation over the white matter after fixation in 4% parapharmaldehyde solution for six hours then in protective solution 30% glucose for one week,then under examination multiple punctate hemorrhages visible in the red circles in picture.

### Microscopic observations

#### Haematoxyllin and eosin statining

In 5 out of 7 cases, H&E staining showed destruction of neurons and axons architectures, appearing black in color under the microscope. We also observed hollow formations ballooning in and around the nuclear material and clear vacuolation around the axon (Fig. 2A). Focal, localized hemorrhage was observed in the white matter of the brain in electrocution deaths (Fig. 2C).

**Figure 02:**
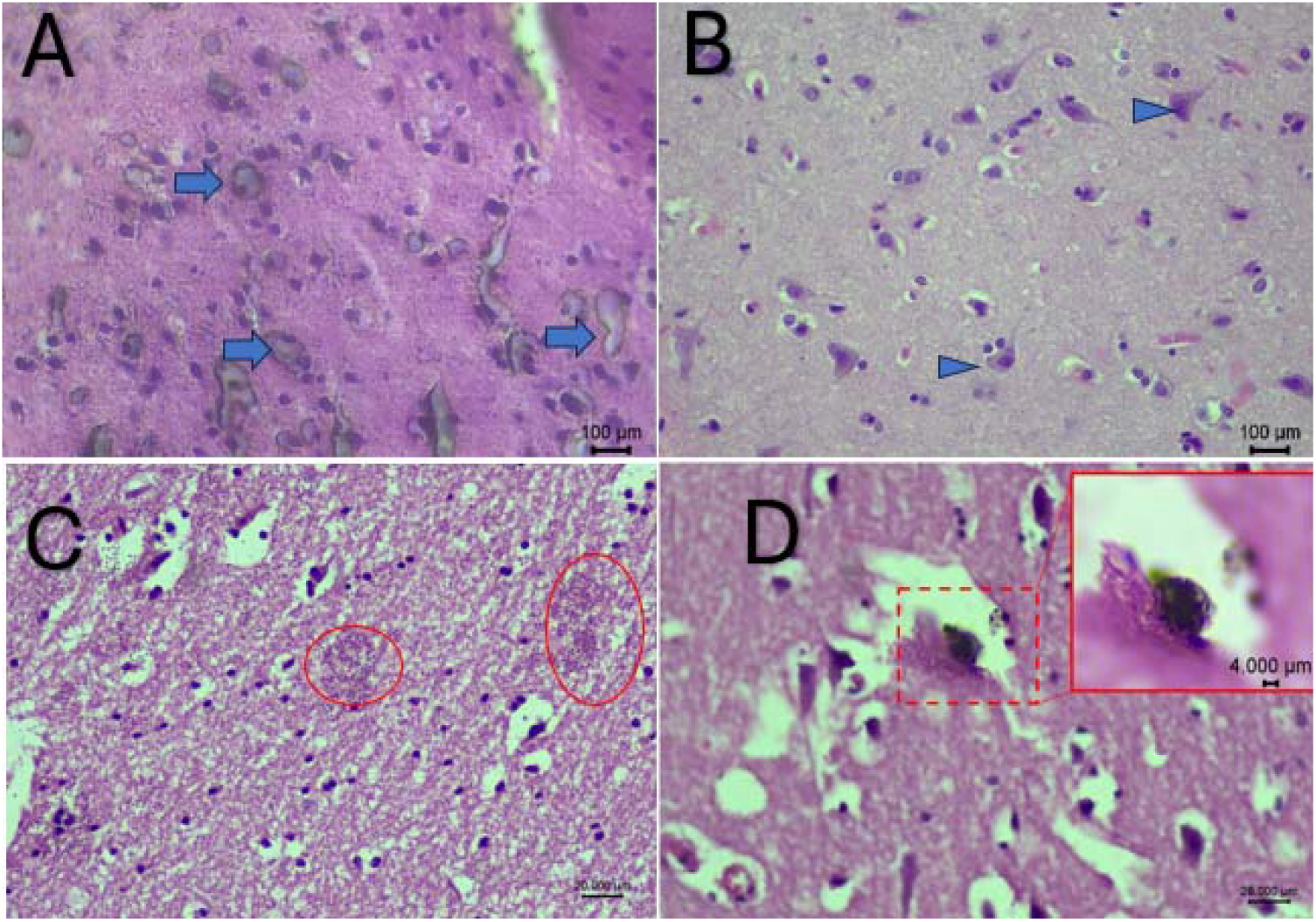
Haematoxyllin and eosin (H&E) statining of the conrtical tissue death due to electrocution. A: Showing distruction of neurons and axons by endogenous heat completally lost the anatomical strucre (Blue arrow head). B: Normal neuronal cell body where deceased died due to unknown poisoning substances(non electrocution death), pyramidal structure of neuronal cell body maintained (Blue arrow head). C:Showing focal heamorrhages at places in the cortical brain tissue electrocution death (Red circle). D:Showing distruction of focal neuronal neuronal material with fragmentation (Red inset).

### Golg cox staining

In Golgi-Cox stained sections, neuronal degeneration was evident, characterized by destruction of neurons, dendrites, and neuronal cell bodies, along with fragmentation of axons and loss of dendritic arborization. The surfaces of these structures appeared irregular, pyramidal neurons had lost their characteristic pyramidal shape and were transformed into rounded or circular forms (Fig. 3B,C). In advanced stages of degeneration, nuclear material appeared aggregated into irregular masses, and in some areas formed clustered, bunch-ofgrapes-like structures (Fig.3 B,C). These features were observed in both the cerebral cortex and cerebellar neuronal tissue. In control cases, the normal structure of neuronal neuropil is appreciable, showing pyramidal-shaped normal architecture neurons. Golgi staining revealed normal architecture of the neuronal cell body and normal architecture of neuronal arborization (Fig.3A).

**Figure 03:**
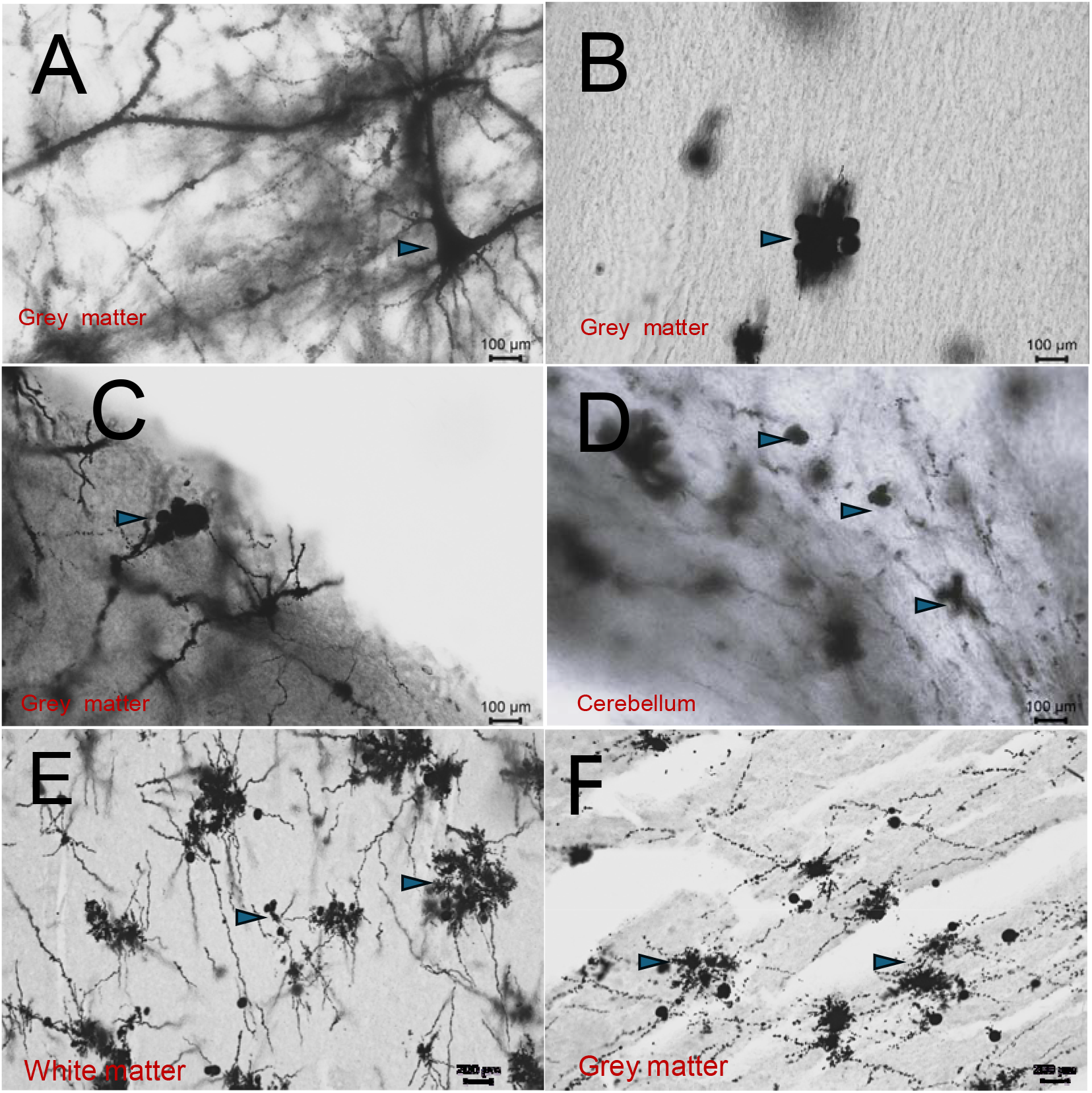
Golgi -cox staining cortical and cerebellum tissue in electrocution death A: Nornal dendritic arboriation and triangular oyramidal shaped neuronal cell body with intact axon death due to hanging (non electrocution death). (B&C): Ballooning and agregation of neuronal material looking like bunch of grapes like structure in the cortical tissue. D: In celebellum agrregationand distruction of neuronal cells, E&F: In white and grey mater fragmentation in multiple small ball like structure of neuronal material distruction and alteration of the normal architectural arboriztion, all this distruction due to the effect of endogenous heat, produced and travels through the nerve fibres in the electrocution death.

**Table 2:** Observations.

|  | Focal haemorrhage n-7 | Axonal vacuolation n-7 | Neuronal destruction n-7 |
| --- | --- | --- | --- |
| 1 | 4 | 5 | 5 |

These finding (focal haemorrhage, axonal vacuolations and neuronal distruction) were not seen in control cases as taken as asphyxia deaths,poisoning cases. Time since death reported all cases between 6-18 hours.

### Statistical analysis

To evaluate neuronal size in electrocution and non-electrocution deaths, histological sections of the brain were examined under a light microscope. In each case, five representative microscopic fields were selected. Within each field, the area of all identifiable neurons was measured using the freely available Image J software (National Institutes of Health, USA). (steps-selection the picture-set mearurents -image change in 8 bitsthresolding-anylize the particle size 50-infinite in micron, then final summery result obtained). The average neuronal area for each case was then calculated. The same methodology was applied to the non-electrocution (non electrocution death) cases, and the mean neuronal areas of the two groups were compared.

Similarly, the cross-sectional area of transversely cut blood vessels was measured in five representative microscopic fields from each case using Image J software. The average vessel area was calculated for each case and compared between electrocution and non-electrocution deaths and statistically analyzed. The statistical significance was set at p ≤ 0.05. All statistical analysis were performed using jamovi version 2.8.1.

**Figure 4:**
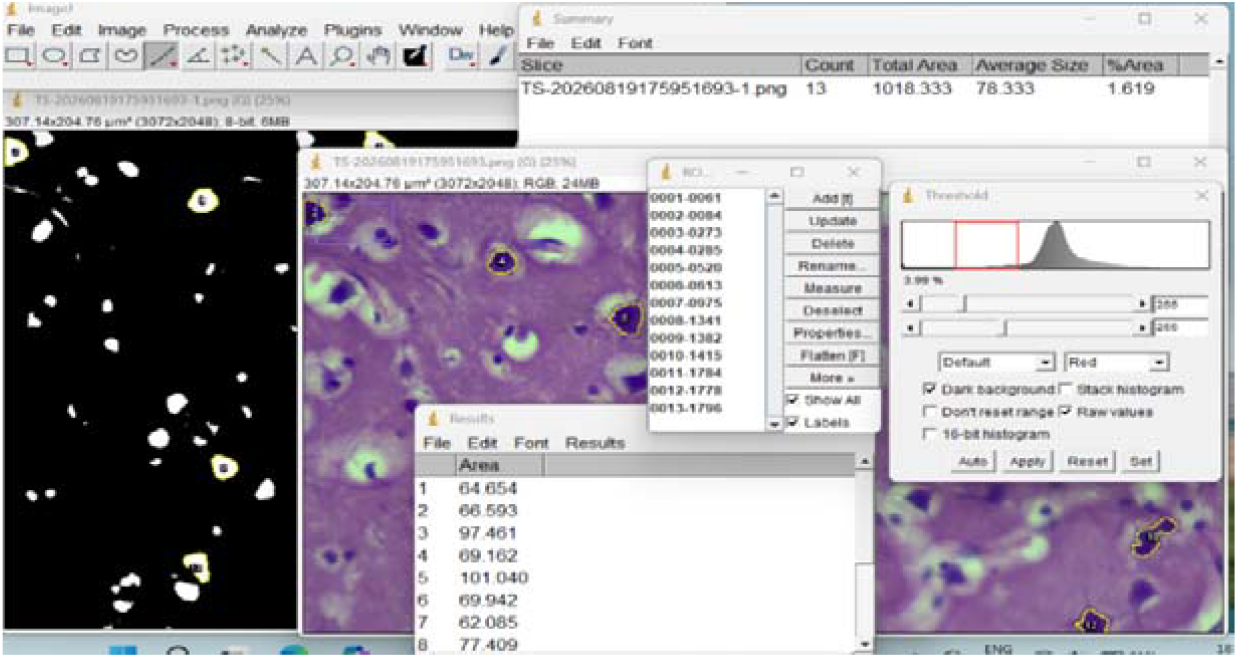
Photographic detail of method used for the cross section area measurment of neuron/vessels by national institute of health USA (Imaje j shoftware).

The mean neuronal cross-sectional area was 84.86 +10.54µm^2^ in the electrocution group and 80.86 +12.15µm^2^ in the control group. The mean difference was 4.00µm^2^, corresponding to an approximately 4.95% higher mean value in the electrocution group. Welch’s independent-samples - test showed no statistically significant difference between the groups,t (7.93)= 0.594, p=0.569,95% CI (−11.56 -19.56) The Mann–Whitney U test produced a similar result,U=23.0,p=0.432. Cohen’s d was 0.357, indicating a small-to-moderate observed effect (fig. 5).

**Figure 5:**
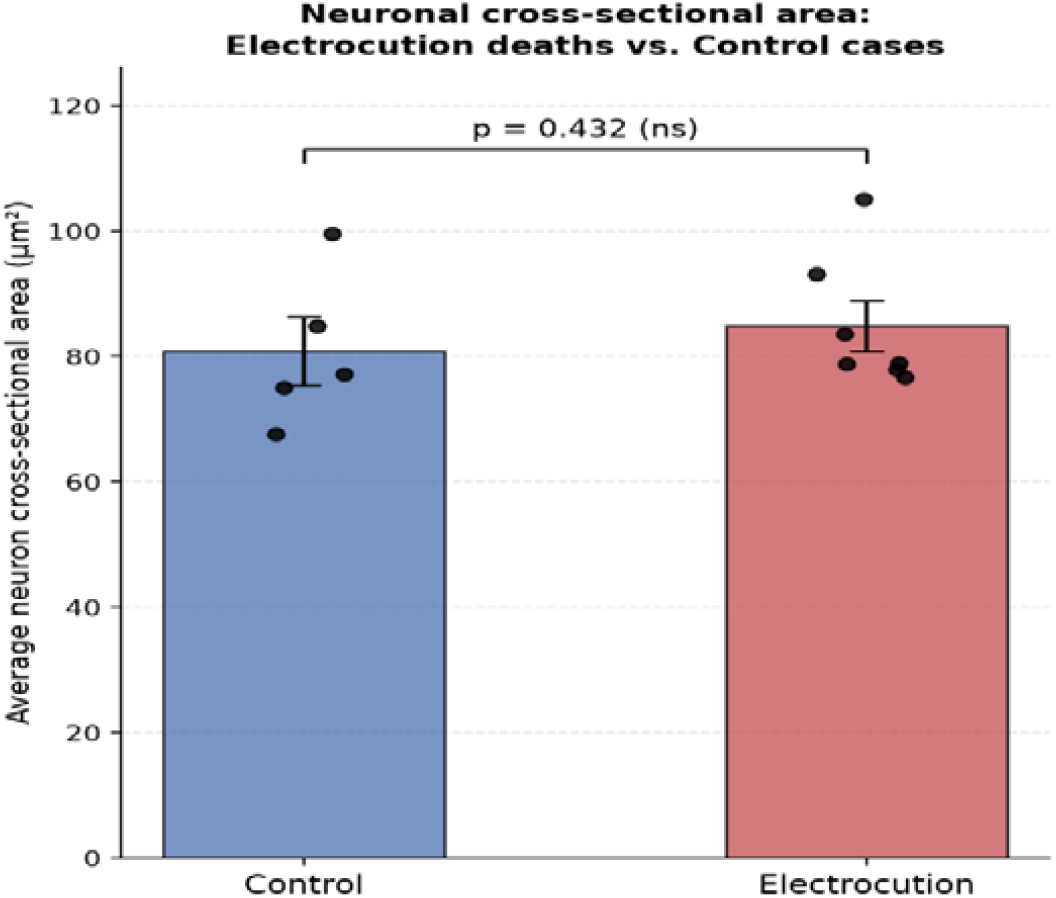
Comparison of average neuronal area (µm^2^) between electrocution and non-electrocution eaths.

The mean cross-sectional vessel area in electrocution deaths (86.74 ± 63.40 µm^2^, n = 7) was approximately 2.6-fold higher than in control cases (33.48 ± 30.25 µm^2^, n = 5). Given nonnormal distribution in the control group (Shapiro-Wilk p = 0.010), a Mann-Whitney U test was applied, showing a statistically significant difference between groups (U = 30.0, p = 0.048). The effect size was large (Cohen’s d = 1.01), supporting vascular dilation/congestion as a histomorphological correlate of electrical injury (fig.6).

**Figure 6:**
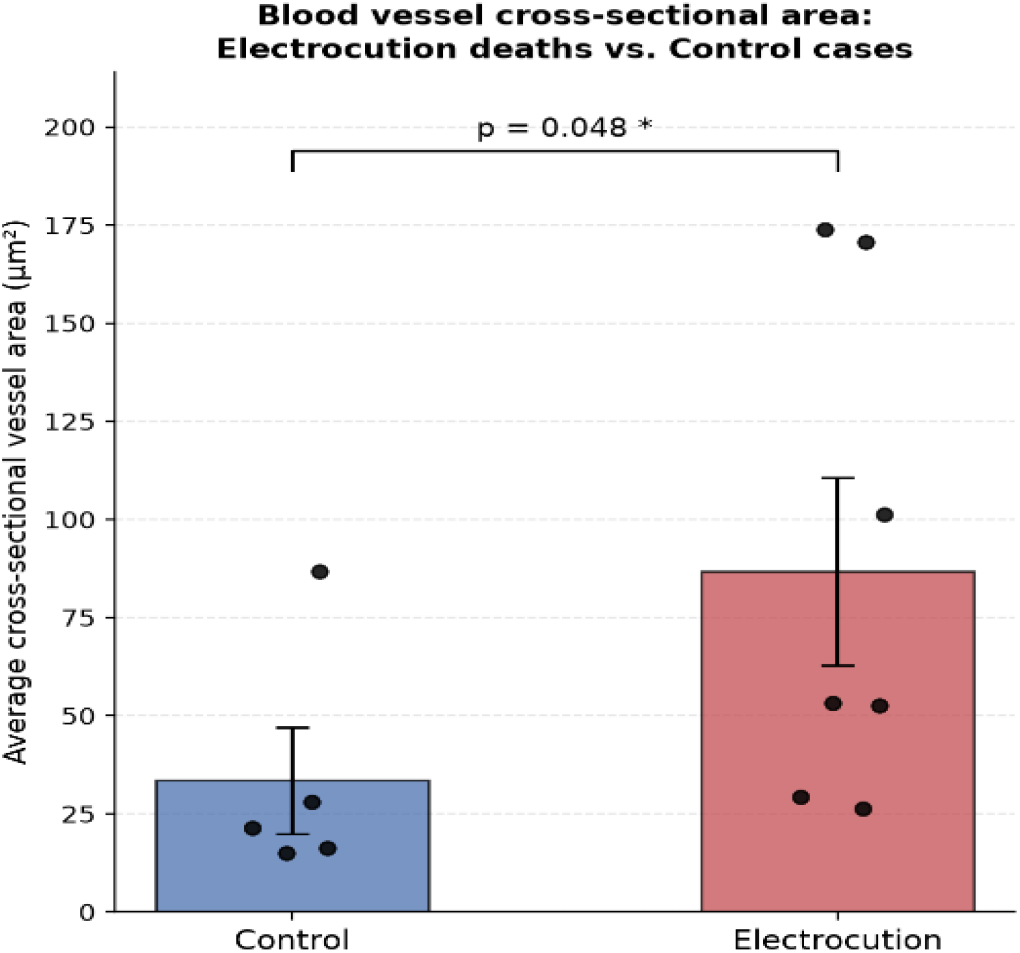
Comparison of mean vessel dilatation area (µm^2^) between electrocution and nonelectrocution deaths.

## Discussion

The results of this study are significant and demonstrated distinctive diagnostic value that changes in neuronal tissue following the passage of household electrical current (220–240V). Amount of current generating the neuronal circuit in nornal functioning condition is relatively very small, the normal neuronal resting membrane potential (−70mV to -80mV) and maximum action potential range (+100 mV), the voltage difference More than 200 mV cause lectrical breakdown, thereby surpassing tissue tolerance thresholds (7). This exposure produces characteristic morphological alterations in neurons that may serve as valuable diagnostic indicator for electrocution death, particularly in cases where external electrical marks over the body found absent.

Does electrocution affect a specific part of the brain or the whole brain? Evidences are available the CNS parital, temporal and cerebellum, other part of the CNS also affrected by the endogenous heat where somatocenssory fibers connected with the anterior hypothamas and medulla oblangata(8,9). In high voltage electrocution death, brain hemorrhage,axonal balloon degeneration in white matter necrosis and degeneration are observed (10–12). Axonal fragmentsations are also observed in the animal study (13). The severity of an electric shock varies which based on the current type, intensity, duration, pathway, location of damage, and physical conditions(14). Neuronal changes seen in Golgi cox staining is not due to decomposition as there is liqufactive necrosis seen after death (15). In the daily practice of the forensic pathologist, electrocution death is o ne of the most difficult diagnosis (10). The main reason of this difficulty is the lack of any specific internal finding. Cause of death in electrocution mainly decided by entry and exit wound over the external surface of body (5).

Most common cause of death is ventricular fibrillation in electrocution death which can not be diagnosed by autopsy examination or by any investigation (16). As presented in Table 01, neuronal alterations in brain tissue were found to be independent of the direct passage of electrical current through the brain. Notably, even when the current pathway was not passed through the brain pathway it observed the electric current pathway between the entry and exit sites (e.g. from the right hand to the right foot) as significant morphological changes were observed in the brain tissue. Our histopathological observation shown destruction of neurons ,ballooning of axons specific indicative for the electrocution. Electricity induced Purkinje cell apoptosis, hallowed with degeneration seen in rat study with shrunken, irregular outline purkinge cells indicates death due to electrocution (17). Our study also observed Purkinje cell, and other nuclear material destruction in cerebellum.

Electron microscopy also indicated nuclear irregularities with nuclear indentation, there is a massive accumulation of cytoskeletal elements in its perikaryal by silver staining, perinuclear space electrocution (17). CT scan finding in electrocution bilateral cerebellar and left occipital hypodensity and thalamic bleed seen in electrocution (18–20). Electrocution remains one of the most difficult diagnoses in forensic pathology due to the lack of consistent internal findings. Absent of external electrical marks with cardiac arrhythmias causing death without structural trace.

In this study we demonstrated that neuropathological examination can provide valuable diagnostic clues. Neuronal destruction and axonal ballooning observed that indicate electrical injury to neural tissue. These findings are consistent with experimental animal studies demonstrating neuronal damage following electrical exposure. Cerebellar involvement, particularly Purkinje cell loss is noteworthy. Previous animal studies have shown Purkinje cell apoptosis following electrical injuries, Pyramidal cell loss and reduction in numbers (21,22), leptomeningeal hemorrhages and disruptions (23), also neuronal loss in the spinal cord (24). supporting the findings of the present study.

Notably, these histopathological findings were not consistently present across all samples. Furthermore, even within individual specimens, features suggestive of electrocution were observed to be heterogeneously distributed. Golgi–Cox staining provided additional insights into dendritic arborization damage, which is not easily and confirmatory appreciable with routine H&E staining. This highlights its importance as an adjunct technique in forensic neuropathology. Observed findings along with supportive circumstantial evidence, can strengthen the diagnosis of electrocution death.

## Future Directions

Future studies with larger sample sizes and incorporation of immunohistochemical markers (e.g., apoptosis and heat shock proteins) are recommended to further elucidate the mechanisms of electrical injury.

## Conclusion

Neuropathological examination significant neuronal and dendritic alterations in fatal electrocution. These findings can serve as supportive diagnostic markers in determining cause of death, particularly in cases lacking external electrical injuries.

## Data Availability

All data produced in the present work are contained in the manuscript.

## CRediT authorship contribution statement

AKR: Conceptualization, data curation, formal analysis, investigation, methodology, supervision, writing original draft, writing review & editing; AK&RK : data curation, formal analysis, investigation; YMR&DDN : Sample collection and laboratory support for Golgi analysis and photograph capturing, SKP&TDW: supervision, writing-review & editing. MDD: Writing-review & editing.

## Funding statment

We did not receive any specific grant from funding agencies in the public, commercial, or not-for-profit sectors.

## Ethical statements

Institutional ethical clearance obtained as IEC no. AIIMS/Pat /IEC/1163/23.

## Acknowledgments

The authors acknowledge the support of Dr. Amit M Patil, Head of the Department of Forensic Medicine, AIIMS Patna, for providing departmental help and institutional facilities. They also thank the laboratory technical staff for their assistance in conducting this study.

## References

1. Dettmeyer RB, Verhoff MA, Schütz HF. Forensic medicine: fundamentals and perspectives. Springer Science & Business Media; 2013 Oct 9.

2. Kuhtic I, Bakovic M, Mayer D, Strinovic D, Petrovecki V. Electrical mark in electrocution deaths–a 20-years study. Open Forensic Sci J. 2012;5(1):23–7.

3. Irasci Y, Goren S, Subasi M, et al. Electrocution-relatedmortality: a review of 123 deaths in Diyarbakir, Turkeybetween 1996 and 2002.Tohoku J Exp Med 2006; 208:141–145.

4. Wright RK and Davis JK. The investigation of electricaldeaths: a report of 220 fatalities. J Forensic Sci 1980; 25:514–521.

5. Reddy KN and Murthy OP. The essentials of Forensic Medicine & Toxicology. 35th ed. Jaypee; 2022. 254–256.

6. Zaqout S, Kaindl AM. Golgi-Cox staining step by step. Frontiers in neuroanatomy. 2016 Mar 31;10:38.

7. Zhang J. Basic Neural Units of the Brain: Neurons, Synapses and Action Potential. 2019;1–38.

8. Purves D, Augustine GJ, Fitzpatrick D, Katz LC, LaMantia AS, McNamara JO, Williams SM. The major afferent pathway for mechanosensory information: the dorsal column-medial lemniscus system.Neuroscience. 2001.

9. Al-Chalabi M, Reddy V, Alsalman I. Neuroanatomy, posterior column (dorsal column). 2018;

10. Visonà SD, Chen Y, Bernardi P, Andrello L, Osculati A. Diagnosis of electrocution: the application of scanning electron microscope and energy-dispersive X-ray spectroscopy in five cases. Forensic science international. 2018 Mar 1;284:107–16.

11. Schulze C, Peters M, Baumgärtner W, Wohlsein P. Electrical Injuries in Animals: Causes, Pathogenesis, and Morphological Findings. Vet Pathol. 2016;53(5):1018–29.

12. Tanda E, Genadiev G, Zappadu S, De Donno G, Serra A, Lamia I, et al. Electrocution as a cause of vascular injury: Case series and literature review. Ann Vasc Surg -Br Reports Innov. 2023;3(2):100201.

13. Mansueto G, Di Napoli M, Mascolo P, Carfora A, Zangani P, Pietra B Della, et al. Electrocution stigmas in organ damage: The pathological marks. Diagnostics. 2021;11(4).

14. Duff K, McCaffrey RJ. Electrical injury and lightning injury: A review of their mechanisms and neuropsychological, psychiatric, and neurological sequelae. Neuropsychol Rev. 2001;11(2):101–16.

15. Krassner MM, Kauffman J, Sowa A, Cialowicz K, Walsh S, Farrell K, et al. Postmortem changes in brain cell structure: a review. 2023;10:1–53.

16. Shaha KK, Joe AE. Electrocution-related mortality: a retrospective review of 118 deaths in Coimbatore, India, between January 2002 and December 2006. Medicine, Science and the Law. 2010 Apr;50(2):72–4.

17. Kandeel S, Elhosary NM, El-Noor MM, Balaha M. Electric injury-induced Purkinje cell apoptosis in rat cerebellum: histological and immunohistochemical study. Journal of chemical neuroanatomy. 2017 Apr 1;81:87–96.

18. Jang SJ, Wi DH. Right Thalamic Hemorrhage Resulting from Low-voltage Electrical Injury: A Case Report. Journal of The Korean Society of Emergency Medicine. 2012 Aug 1;23(4):555–8.

19. Çaksen H, Yuca SA, Demirtas I, Odabas D, Cesur Y, Demirok A. Right thalamic hemorrhage resulting from high-voltage electrical injury: a case report. Brain and Development. 2004 Mar 1;26(2):134–6.

20. R. Singh Jain, S. Kumar, D. T. Suresh, and R. Agarwal, “Acute vertebrobasilar ischemic stroke due to electric injury,” The American Journal ofEmergencyMedicine,vol.33, no.7,pp. 992–992.e6, 2015.

21. Huang, Q. Y., Chen, Y. C., & Liu, S. P. (2012). Reduction in Purkinje fiber number in rats undergone fatal electrocution. The American Journal of Forensic Medicine and Pathology, 33(1), 19–21.

22. Kurtulus A, Acar K, Adiguzel E, Boz B. Hippocampal neuron loss due to electric injury in rats: A stereological study. Leg Med. 2009;11(2):59–63.

23. Sancesario G, Massa R, Petrillo S, Nottola SA, Correr S, Rossini PM. Transcranial unifocal stimulation in rabbit: subcutaneous and meningeal changes. European neurology. 1989 Feb 6;29(2):93–8.

24. Seo CH, Jeong JH, Lee DH, Kang TC, Jin ES, Lee DH, Jeon SR, Choi KH, Hwang HS. Radiological and pathological evaluation of the spinal cord in a rat model of electrical injury-induced myelopathy. Burns. 2012 Nov 1;38(7):1066–71.

